# Potential impact of a population-based screening program on the increased burden of prostate cancer in Thailand: A simulation study

**DOI:** 10.1101/2024.10.29.24316321

**Authors:** Christian S. Alvarez, Alison M. Mondul, Laura S. Rozek, Hutcha Sriplung, Rafael Meza, Jihyoun Jeon

**Affiliations:** Department of Epidemiology, University of Michigan School of Public Health, Ann Arbor, MI, U.S.A; Department of Oncology, Georgetown University Medical Center, Washington D.C., U.S.A; Epidemiology Unit, Prince of Songkla University, Faculty of Medicine Hat Yai, Hat Yai District, Songkhla, Thailand; Department of Integrative Oncology, BC Cancer Research Institute, Vancouver, British Columbia, Canada

**Keywords:** Prostatic neoplasms, Screening, Thailand, Simulation

## Abstract

**Background:** Prostate cancer incidence and mortality are expected to increase considerably in the near future in Thailand. There is thus an urgent need to establish prevention measures, such as screening, to reduce the increasing burden of prostate cancer in Thailand.

**Methods:** Using data from several sources including the Songkhla Cancer Registry and the census data from Thailand, we conducted a simulation analysis to assess the potential impact of screening on the incidence and mortality of prostate cancer among 10 million males aged 50 to 70 of 1960 birth cohort from Songkhla, Thailand. We assumed 4 different scenarios, including no screening, 15%, 60% and 100% screening uptakes of the prostate-specific antigen test. Furthermore, stage distribution of prostate cancer was assumed based on two major prostate cancer screening trials: European Randomized Study of Screening for Prostate Cancer (ERSPC) and Prostate, Lung, Colorectal, and Ovarian (PLCO) Cancer Screening Trial. The number of prostate cancer cases was projected using an age-period-cohort model approach, accounting for the expected excess of cases due to screening. Deaths from prostate cancer were then projected using survival probabilities from Songkhla and the United States. Case fatality ratios (CFRs) were also computed.

**Results:** Prostate cancer incidence increased with screening, as expected, with a shift of the stage distribution toward earlier stages, but mortality from prostate cancer decreased with higher screening uptake. Assuming 1.71 excess risk of cases due to screening and stage distribution from the ERSPC trial, we projected an increase of over 7,000 localized cases under 100% screening uptake, while the cases in advanced stages decreased from 4,046 (no screening) to 96 under 100% screening uptake. The number of deaths were reduced by 82% under 100% screening uptake compared to no screening. The CFR also decreased from 0.42 (no screening) to 0.05 (100% screening).

**Conclusion:** Screening for prostate cancer could substantially reduce the number of prostate cancer cases in advanced stages and prostate cancer deaths. Although the net benefit depends on the assumed survival rates under screening, which could vary depending on the quality of the implementation, screening would contribute to reducing the escalating burden of prostate cancer in Thai population.

## Introduction

Prostate cancer is emerging as a significant public health problem in many developing countries.^1–3^ In Thailand, there has been a significant increase in the incidence and mortality rates of prostate cancer over the last few decades,^4–6^ with a large proportion of prostate cancer cases diagnosed at advanced stages. In Songkhla province, approximately 75% of staged tumors are stage IV at diagnosis.^7^ In contrast, in the United States (US), the vast majority of prostate cancer cases are diagnosed at early stages (74% localized),^8^ which is partially explained by the widespread use of the prostate-specific antigen (PSA) test for prostate cancer screening in the US population.^9^ Rates of PSA testing in the US have recently increased after reversing previous national guidelines on recommending against PSA testing for all individuals.^10^

Screening for prostate cancer is controversial as it leads to a considerable increase in incidence while the net benefit for prostate-specific mortality remains unclear.^11^ Two major randomized clinical trials have been conducted to assess the effectiveness of PSA screening in the reduction of prostate cancer mortality producing conflicting results.^12,13^ The European Randomized Study of Screening for Prostate Cancer (ERSPC) conducted in several European countries showed a statistically significant reduction in prostate cancer mortality of 20% (Rate Ratio [RR]: 0.80; 95%CI: 0.72, 0.89) among men who underwent PSA screening after 16-years of follow-up.^13^ Conversely, the Prostate, Lung, Colorectal, and Ovarian (PLCO) Cancer Screening Trial in the US showed no statistically-significant reduction in prostate cancer mortality among men who were screened during the same follow-up period (RR: 0.93; 95%CI: 0.81, 1.08).^12^ The discrepancy between these trials has been largely attributed to screening occurring outside of the trial in the placebo arm of the PLCO trial (contamination).^14^ Despite the critical need for early detection of prostate cancer, PSA screening is also associated with potential harm as a result of overdiagnosis and subsequent overtreatment, which leads to adverse effects, particularly in older men.^15^ Thus the adoption of PSA screening in other countries, such as Thailand, remains controversial.

Currently, there is no population-based screening program for prostate cancer in Thailand, where the burden of the disease is expected to continue to rise.^6^ Therefore, assessing the impact of screening strategies for the control of prostate cancer is necessary for this country. We, therefore, conducted a simulation analysis to evaluate the potential impact of screening, using the PSA test on the incidence and mortality of prostate cancer, while taking into account the potential downsides from overdiagnosis.

## Methods

### Data sources

Incident prostate cancer cases from the Songkhla Cancer Registry and census data from the National Statistical Office of Thailand were used in this analysis. The Songkhla Cancer Registry collects information on age, year of diagnosis, religion, stage, and grade, as well as the date of last contact, date of death, and vital status. A total of 946 prostate cancer cases were diagnosed in Songkhla, Thailand between 1990 and 2014, with all-cause mortality of 61.9% during the same period. We obtained population denominators from decennial census data in 1990, 2000, and 2010. The annual intercensal population structure in Songkhla was obtained by 1-year for each sex-specific group. The population for the Songkhla province beyond 2010 was estimated by the Thai Office of the National Economic and Social Development Board. Our simulation analysis included population data from Songkhla males up to 2030. This study is a secondary analysis of de-identified cancer registry data and thus the institutional review board approval is not required.

### Screening scenarios

We divided the Songkhla male 1960 birth cohort population (ages 50 to 70 years old), into screened and unscreened individuals assuming the following screening uptake rates: 100%, 60%, and 15%. The last two screening scenarios are based on reports of the prevalence of PSA screening in the US and other Asian countries. For example, in 2010, the Behavioral Risk Factor Surveillance System (BRFSS) —a nationally representative survey on health-related risk behaviors and use of preventive services in the US,^16^ reported that approximately 60% of US males aged 50 years and older had undergone PSA screening for prostate cancer in many states.^16^ On the other hand, a Japanese study reported a PSA screening prevalence of 12% in males aged 55-69 from Kanazawa City in 2010.^17^ In addition, a Chinese study reported a 10% prevalence of PSA screening among males aged 50 and older.^18^ We compared the results based on these screening scenarios with no screening.

Our target population was men from Songkhla born in 1960. We followed this cohort from ages 50 to 70, which spans years 2010-2030. We started at age 50 because of the recommendations in existing prostate cancer screening guidelines, such as, the Japanese Urologic Association (50 years and older), the PLCO trial (50 to 74 years), the ERSPC trial (55 to 74 years), and the US Preventive Services Task Force (USPSTF) (55 to 69). On the other hand, the USPSTF recommends against the use of PSA screening for prostate cancer in males aged 70 and older.^19^ Therefore, we truncated our target population at age 70.

### Simulation analysis

#### Projections of prostate cancer

We simulated 10 million individuals aged 50-70 in birth cohort 1960 from the Songkhla male population. To project the number of prostate cancer cases under different screening scenarios, first we used the population incidence rates of prostate cancer in Songkhla. The prostate cancer rates were modified to account for the excess of cases among screened population. Therefore, the prostate cancer rates were multiplied in the baseline scenario by 1.71, which reflects the ∼70% excess of cases under PSA screening observed in the ERSPC at the beginning of the trial;^13^ in addition, rates were multiplied by 2.0 and 1.5 as upper and lower bound for the excess of cases (results from both scenarios are presented as supplemental materials). The prostate cancer incidence rates were computed by age and period using a previously developed age-period-cohort model^20^ based on data from the Songkhla Cancer Registry: For age a and calendar year p, the prostate cancer incidence rate was computed by:

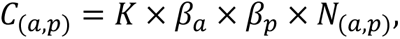

where *β*_*a*_ and *β*_*p*_ are coefficients for age effects and the period effects, respectively, and *N*_(*a*,*p*)_ represents the size of the population at age *a* and calendar year *p*. *K* is an adjusting factor related to screening, which was set to either 2.0, 1.71, 1.5 for those undergoing screening, and 1 otherwise (unscreened).

For the stage distribution of prostate cancer cases among the screened population, we applied the stage distribution of prostate cancer observed in the ERSPC^13^ and PLCO^12^ to the projected cases under the different screening uptake rates. For the distribution of prostate cancer cases under the no screening scenario, we used the observed stage distribution from Songkhla province. The stages of prostate cancer were classified as: localized, regional, and distant.

#### Projection of prostate cancer deaths

To project the number of prostate cancer deaths without screening, we first fitted Weibull survival models to obtain annual survival probabilities by tumor stage (localized, regional, and distant) from the Songkhla Cancer Registry data. Since the prostate cancer specific mortality data was not available in the Songkhla Cancer Registry, we assumed that all-cause mortality among prostate cancer cases in this registry is equivalent to prostate cancer specific mortality in this simulation analysis. We then used the Weibull survival models to project the number of prostate cancer deaths by year among unscreened cases. Because of the large number of unstaged tumors (76%) in the Songkhla Cancer Registry, we used a multiple imputation analysis with chained equations to impute the missing information for the tumor stage.^21,22^ Two parameters were obtained from the Weibull survival model: scale and shape (or slope).^23^ The model adequacy was assessed by inspecting empirical-based Kaplan-Meier curves vs. model estimates (Supplementary Figure 1).

To project deaths from the screen-detected prostate cancer cases, we instead used survival probabilities from Surveillance, Epidemiology, and End Results Program (SEER) 9 registries for US males aged 50 and older from 1990 to 2014, ^24^ representing prostate cancer survival during the PSA screening era. Those survival probabilities were applied to the cases among screened individuals to account for the benefit of screening, assuming that the quality of care would be similar in Thailand to that received by males in the US.

The total number of deaths from prostate cancer between age 50 and 70 for the 1960 birth cohort is calculated by:

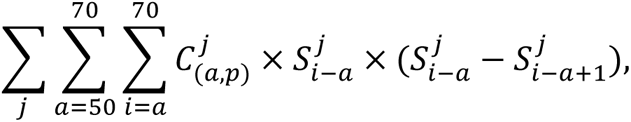

where j corresponds to the tumor stage (localized, regional, and distant), C(a,p) is the number of prostate cancer cases at age a and calendar year p, and S_t_ corresponds to the survival probability at time t. R statistical software (Survival package) was used for this analysis.

We computed case fatality ratios (CFRs) to estimate the overall impact of prostate cancer screening on prostate cancer mortality across screening scenarios. The CFR was calculated by dividing the total number of deaths by the total number of prostate cancer cases. In order to take into account overdiagnosis in the CFR calculations, we removed either 23% (lower bound) or 42% (upper bound) of prostate cancer cases that were screen-detected, because this range corresponds to the overdiagnosis rates observed in the US during the PSA screening era.^25^

Supplementary Figure 2 shows the flowchart for the procedures used in this simulation analysis.

## Results

Of the 946 prostate cancer cases in Songkhla province, 89.2% were Buddhists and the rest Muslims. Over 60% of cases were older than 60 years of age. Of tumors with known grade at diagnosis the majority were moderately/poorly differentiated. Similarly, among tumors with known stage at diagnosis, the majority were distant (**Table 1**).

**Table 1.** Characteristics of cases with prostate cancer (N=946), 1990 to 2014.

| Characteristics | N (%) |
| --- | --- |
| Age at diagnosis, years |  |
| <50 | 5 (0.5) |
| 50-59 | 69 (7.3) |
| 60-69 | 235 (24.9) |
| 70-79 | 407 (43.0) |
| >=80 | 230 (24.3) |
| Year of diagnosis |  |
| 1990-1994 | 87 (9.2) |
| 1995-1999 | 91 (9.6) |
| 2000-2004 | 147 (15.5) |
| 2004-2009 | 265 (28.0) |
| 2010-2014 | 356 (37.7) |
| Religious group |  |
| Buddhists | 844 (89.2) |
| Muslims | 98 (10.4) |
| Others | 1 (0.1) |
| Unknown | 3 (0.3) |
| Tumor stage |  |
| Localized | 7 (0.7) |
| Regional | 43 (4.5) |
| Distant | 149 (15.8) |
| Unknown | 747 (79.0) |
| Tumor grade |  |
| Well differentiated | 186 (19.7) |
| Moderately differentiated | 100 (10.6) |
| Poorly differentiated | 147 (15.5) |
| Undifferentiated | 69 (7.3) |
| Unknown | 444 (46.9) |

**Table 2** shows the cumulative number of prostate cancer cases and deaths under various screening scenarios with three selected screening coverages (15%, 60%, and 100%). As expected, there is a shift in the stage distribution of prostate cancer towards more localized stages, particularly under 100% and 60% uptake rates. This reflects the difference in the assumed stage distribution between the no screening and the screening scenarios. The simulations show a higher number of stage III in the ERSPC than PLCO trial scenarios, particularly under 100% screening uptake, which reflects the difference in distributions between both trial scenarios. Overall, the number of prostate cancer deaths decreased with higher percentage of screening uptake. For example, with no screening, the overall number of deaths projected was 2,155 (stage I & II: 45; stage III: 289; stage IV: 1,821); while the overall number of deaths projected under 100% screening uptake was 387 (stage I & II: 288; stage III: 48; stage IV: 51) and 425 (stage I & 2: 320; stage III: 8; stage IV: 97) in the ERSPC and PLCO trial scenario, respectively.

**Table 2.**
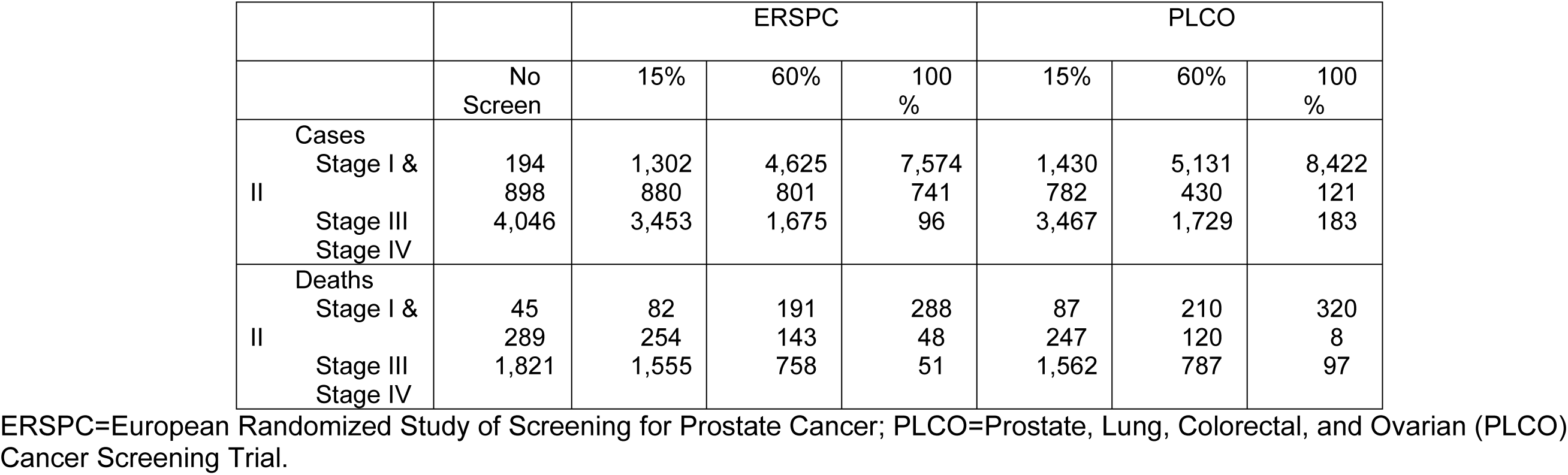
Cumulative number of prostate cancer cases and deaths (ages 50-70) under various screening scenarios with three selected screening coverages (15%, 60%, 100%) in the Thai population.

**Figure 1** illustrates the stage distribution of prostate cancer cases under different screening scenarios. As mentioned above, there is a shift in the stage distribution of prostate cancer cases towards more localized stages. In total, we projected over 7,500 localized cases with only 96 cases with distant stage under 100% screening uptake under the ERSPC trial scenario. In contrast, with no screening, we projected 194 localized cases and 4,046 distant cases. Similar patterns in the stage distribution for prostate cancer as a function of screening coverage are observed under the PLCO trial scenario.

**Figure 1.**
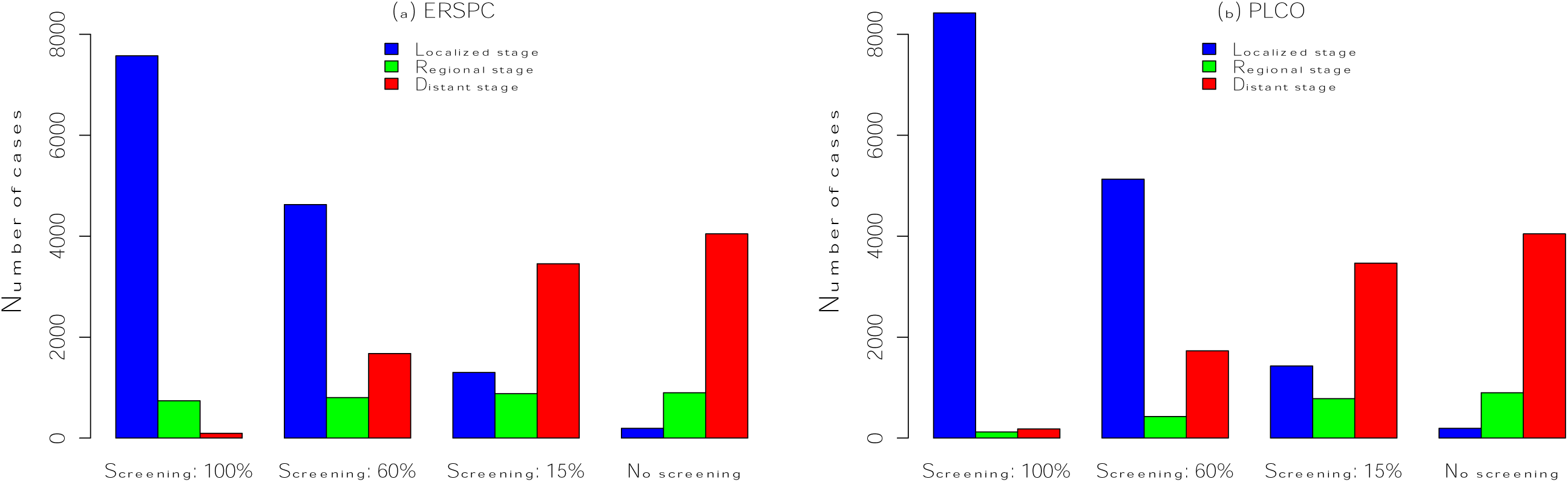
Prostate cancer stage distribution by different screening uptake rates (no screening vs. screening with uptake 15%, 60%, 100%) under the ERSPC and PLCO trial scenarios. ERSPC=European Randomized Study of Screening for Prostate Cancer; PLCO=Prostate, Lung, Colorectal, and Ovarian (PLCO) Cancer Screening Trial.

### Impact of screening on prostate cancer incidence

The prostate cancer incidence increased considerably until 70 years old in our modeled cohort. At age 70, we projected 509 cases and 558 cases in excess under 100% screening uptake compared to no screening (**Figure 2**, **a&c**) under the ERSPC and PLCO trial scenario, respectively.

**Figure 2.**
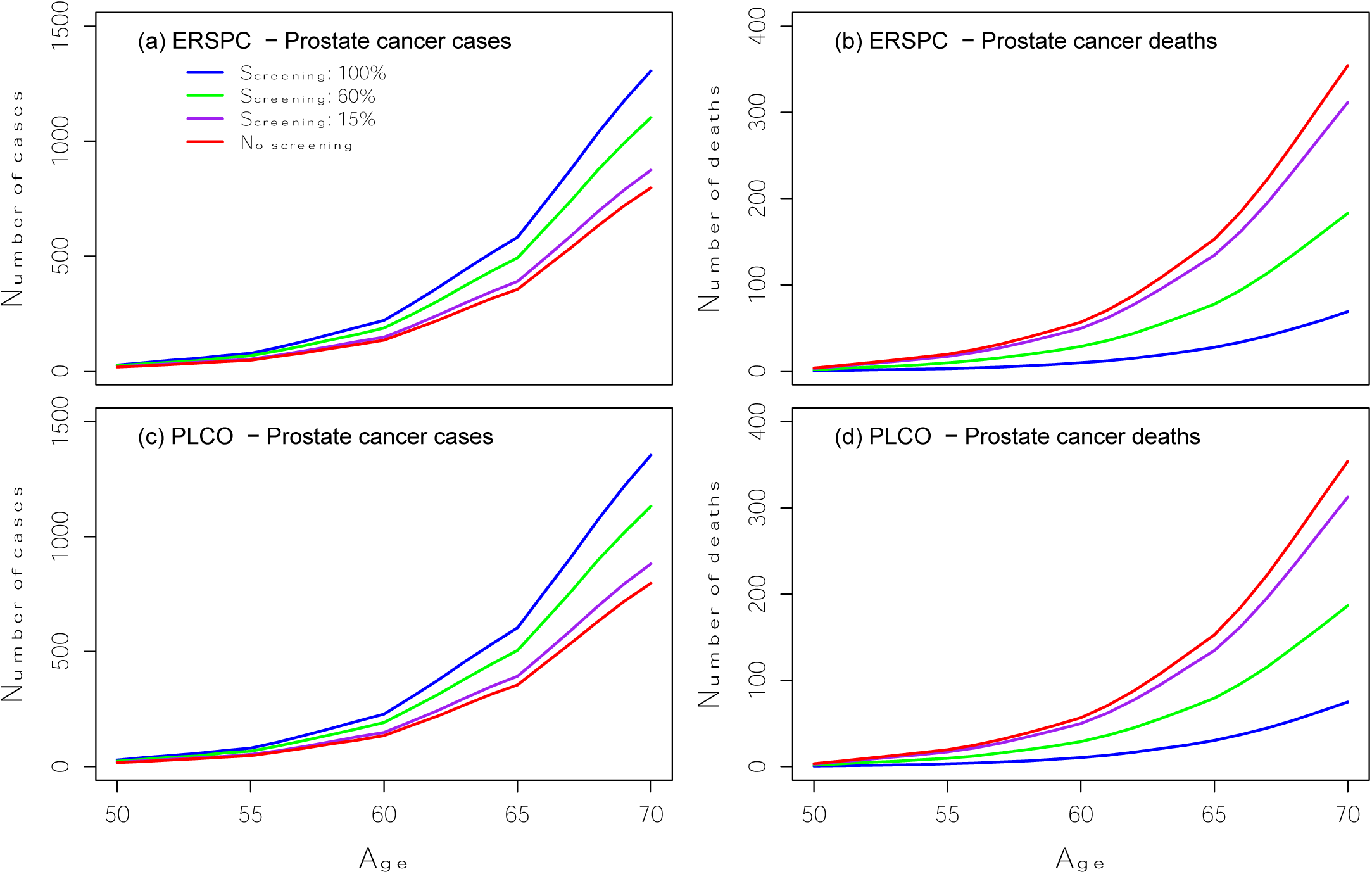
Number of prostate cancer cases and deaths by different screening uptake rates (no screening vs. screening with uptake 15%, 60%, 100%) under the ERSPC (top panels) and PLCO trial scenarios (bottom panels). ERSPC=European Randomized Study of Screening for Prostate Cancer; PLCO=Prostate, Lung, Colorectal, and Ovarian (PLCO) Cancer Screening Trial.

The number of prostate cancer deaths decreased with higher screening uptake (**Figure 2**, **b&d**) under both ERSPC and PLCO trial scenarios. For no screening and 15% screening uptake, the number of deaths is remarkably higher compared to 60% and 100% screening uptake. In total, 1,769 and 1,063 prostate cancer deaths could be prevented with 100% and 60% screening uptake, respectively, under the ERSPC trial scenario. At age 70, the number of prostate cancer deaths decreased by approximately 81% under 100% screening uptake and 49% under 60% screening uptake compared to no screening under the ERSPC trial scenario. Similar reductions were estimated under the PLCO trial scenario.

**Figures 3** shows the number of prostate cancer cases by stage under the ERSPC and PLCO trial scenarios. At age 70, we projected 172, 688, and 1,146 more prostate cancer cases diagnosed at localized stage under 15%, 60%, and 100% uptake rates compared to no screening, respectively, under the ERSPC trial scenario. On the other hand, the number of cases diagnosed at distant stage was greater under no screening compared to any of our modeled screening scenarios over the follow-up time. At age 70, there were 92, 368, and 613 fewer cases with distant stage under 15%, 60%, and 100% screening uptake compared to no screening, respectively, under the ERSPC trial scenario. Similar patterns are observed under the PLCO trial scenario.

**Figure 3.**
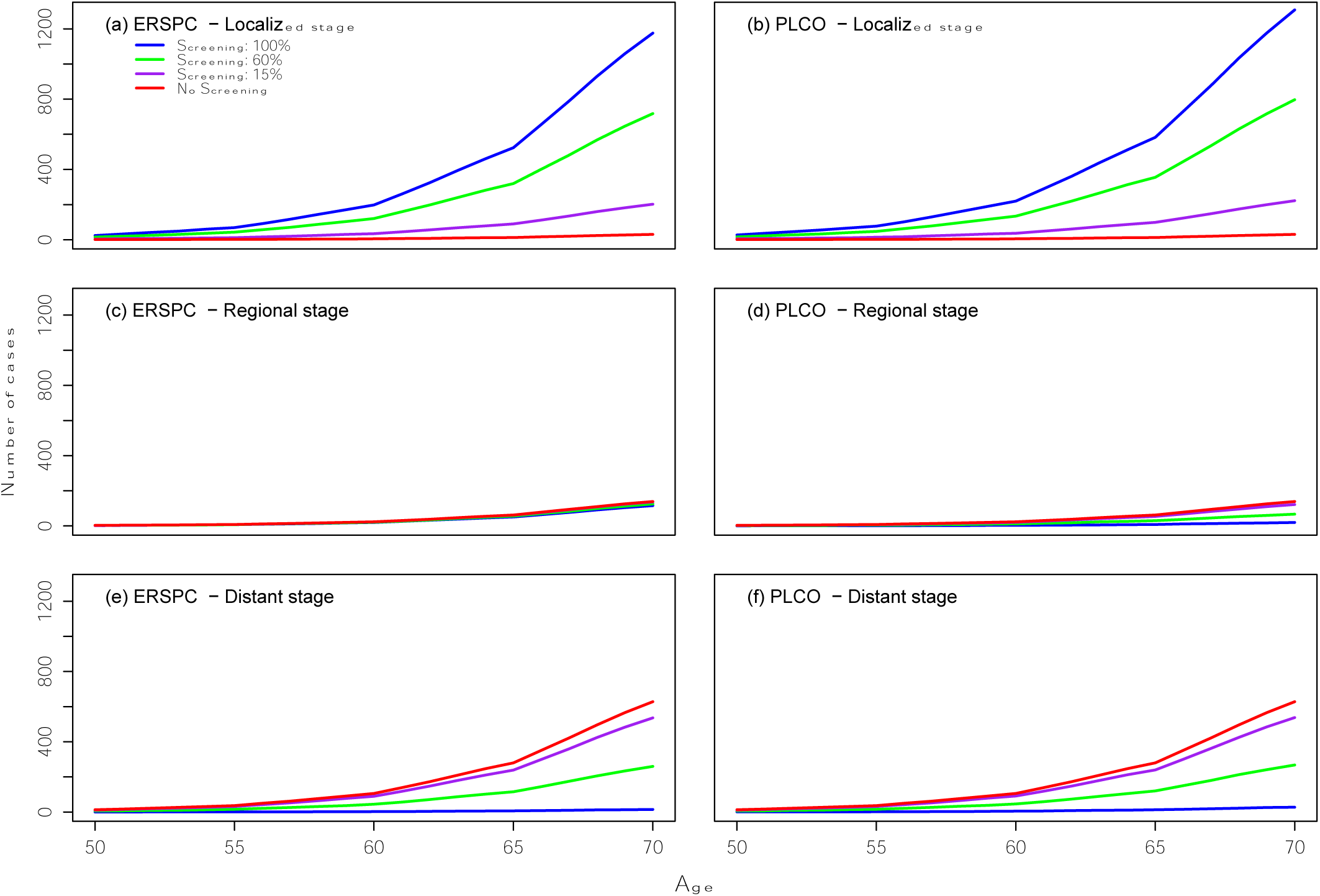
Number of stage-specific prostate cancer cases by different screening uptake rates (no screening vs. screening with uptake 15%, 60%, 100%) under the ERSPC and PLCO trial scenarios. ERSPC=European Randomized Study of Screening for Prostate Cancer; PLCO=Prostate, Lung, Colorectal, and Ovarian (PLCO) Cancer Screening Trial.

**Table 3** shows the case fatality ratios (CFRs) for the different screening scenarios. The CFR decreased from 0.42 with no screening to 0.15 and 0.05 with 60% and 100% screening uptake, respectively, under both ERSPC and PLCO trial scenarios. In addition, under the ERSPC trial scenario, the CFR assuming 23% and 42% of overdiagnosis is approximately 0.18 and 0.20 for 60% screening uptake, respectively. For 100% uptake rate, the CFR assuming 23% and 42% of overdiagnosis is 0.05. These results are similar under the PLCO trial scenario. In general, CFR gets higher with accounting for overdiagnosis.

**Table 3.**
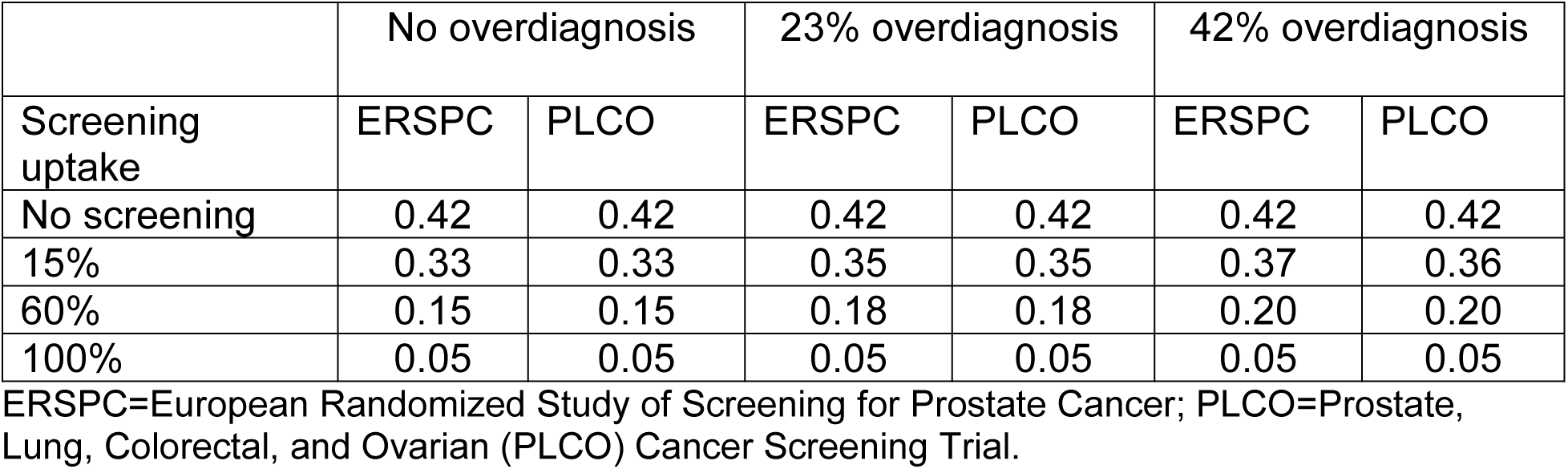
Case fatality ratio (CFR) by different screening uptake rates (no screening vs. screening with uptake 15%, 60%, 100%) under the ERSPC and PLCO trial scenarios.

## Discussion

Our simulation study suggests that screening would shift the stage distribution of prostate cancer at diagnosis toward earlier stages, with the shift being more pronounced with higher screening uptake. Our models projected a remarkable increase in prostate cancer cases among simulated individuals aged 50-70. In contrast, prostate cancer deaths would decrease with higher screening uptakes. Moreover, the projections suggest that the case fatality ratio would also decrease considerably, even while accounting for the expected overdiagnosis due to screening.

Results from our simulation study are consistent with previous literature. In the US, the proportion of males diagnosed with localized prostate cancer increased from 30% to 42% during the earliest period of the PSA era, with the rate of advanced prostate cancer decreasing by 75% between 1989-1992 and 1999-2002.^26^ Similarly, a study from Japan reported that the proportion of metastatic disease decreased with increasing use of PSA screening in a population-based screening cohort.^17^ The study reported a 10% reduction in metastatic disease by increasing the exposure rate for PSA screening from ≤ 10% to ≥ 30.1%.^27^

Our simulation analysis did not consider other screening strategies such as digital rectal examination (DRE). In the US, studies have reported that abnormal findings with DRE are associated with the detection of more clinically significant prostate cancer cases (e.g., high grade disease).^28,29^ Nonetheless, DRE is not recommended as primary screening test for prostate cancer because there is a lack of evidence from randomized controlled trials supporting its efficacy in reducing prostate cancer mortality.^28^ A recent systematic review and meta-analysis concluded that DRE is not recommended to screen for prostate cancer in the primary care setting as there is no evidence supporting its efficacy.^30^

According to our analysis, and our assumptions of excess incidence, stage-shift and prostate cancer survival under screening, the number of prostate cancer deaths in Thailand would decrease considerably with PSA screening. This is consistent with the reduction in prostate cancer mortality observed in the ERSPC trial at 16 years of follow up (Mortality RR: 0.80 [95% CI: 0.72, 0.89]).^13^ On the other hand, no statistically-significant mortality reduction was observed in the PLCO trial (RR: 0.93 [95%CI: 0.81, 1.08]), which has called into question the efficacy of screening on the survival of prostate cancer.^12^ However, several studies have concluded that the use of PSA screening prior to randomization, contamination (subjects in the control arm who received screening), and non-compliance limited the ability to demonstrate the efficacy of screening in the PLCO trial.^14,31–35^

In order to implement a successful screening program in Thailand, there must be the resources available to manage the newly diagnosed cases from the screening program. In addition, it would be important to understand potential barriers that could prevent males from undergoing screening for prostate cancer in this population. Anecdotal evidence from physicians in Thailand indicates that Thai males are embarrassed to talk about urinary problems with providers. In addition, like in other countries, males are less likely to seek healthcare and receive preventive care than females in Thailand. Those factors may prevent them from receiving prostate cancer screening even if it were available. However, the infrastructure already exists within the Thai national healthcare system to provide preventive care and males are already getting some type of health promotion. According to the 2021 National Health and Welfare Survey, 25% of Thai males aged 60 years and older received any health promotion service in the past 12 month.^36^

Several groups in Asia have started the discussion about prostate cancer screening in the region. Since 2010, the Japanese Urological Association recommended the use of PSA screening for males at risk of prostate cancer, explaining the potential risks and benefits of screening.^37^ Despite prostate cancer screening has been recognized as an important need for the control of the disease among Asian countries, there are no official guidelines for prostate cancer in Asian countries, except in Japan.^17,37–40^ In general, the prevalence of prostate cancer screening is very low in Asian countries.^17^ A study conducted in China reported a 10% prevalence of PSA screening among males aged 50 and older; ^18^ the study also suggested that screening recommendations by health care providers was positively associated with the screening uptake.^18^ We hope that our study will advance the evidence necessary to make an informed decision about screening for prostate cancer in the region.

### Strengths and limitations

One of the most important strengths of this study is that we used parsimonious models that allow us to simplify our analysis and approximate real-world conditions. Therefore, they are simpler to translate for healthcare authorities and policymakers, with the purpose of helping them to make an informed decision to plan screening strategies for the control of prostate cancer. Another strength is that we used data from the Songkhla Cancer Registry, which represent well the survival pattern in Songkhla population to project the incidence and mortality of prostate cancer under our simulated screening scenarios for this population.

A limitation of our study is that we used all-cause mortality as a proxy of prostate cancer mortality in the Songkhla Cancer Registry, which may underestimate the survival probabilities computed for the unscreened population. The quality of data from death certificates in Thailand is poor, making determination of cause of death difficult.^41^ However, this approach would be fine for late stage of prostate cancer cases. For example, a recent study reported that 79% of patients diagnosed with metastatic prostate cancer in the U.S. die of prostate cancer within 2 years of diagnosis.^42^ Another important limitation is that we used survival assumptions under PSA screening from the US SEER data that may not be appropriate for the Thai population. This could overestimate the reduction in the number of deaths with screening because the survival probabilities could be higher in the US than in Thailand, in general, due to differences in the healthcare system. However, the majority of studies on prostate cancer screening have been conducted in the Western population, and there is limited data in non-Western countries. Moreover, in contrast with the US, Thailand has universal health care which might reduce issues related to access to care that are experienced in the US, resulting in higher overall survival. Independently, we used the best available evidence to conduct our simulation analysis. Lastly, the CFR estimations may get deflated, providing an overly optimistic impact of screening in the reduction of prostate cancer death if they are not adjusted for overdiagnosis. Therefore, we examined the CFRs adjusted by 23% and 42% of overdiagnosis (Table 3). These rates were chosen because modeling studies in the US showed that the overdiagnosis rate during the time of the introduction of PSA screening was between 23% and 42%.^25^ Further studies are warranted to obtain reliable estimates of overdiagnosis when establishing prostate cancer screening programs in the Thai population, which will be important to evaluate the effectiveness of the screening program.

## Conclusions

Screening for prostate cancer in Thailand could have an important impact on the burden of the disease, diagnosing prostate cancer cases at earlier stages when treatment may be effective. Our study shows that there could be a significant reduction in the number of prostate cancer deaths by implementing a screening program in Thai population, although it is important to take into account any potential risk associated with those screening strategies. Further studies should be conducted to understand the barriers to implementing this strategy in the male population of Thailand, and also the potential benefits and harms of introducing PSA screening in this country given limited resources.

## Data Availability

All data produced in the present study are available upon reasonable request to the authors.

## Acknowledgments

We would like to thank the Songkhla Cancer Registry team for the collection and provision of this data. Dr. Alvarez was supported by the Thomas Francis, Jr Endowment Fund.

## Abbreviations

BRFSS: Behavioral Risk Factor Surveillance System
CFR: Case Fatality Ratios
DRE: Digital Rectal Examination
ERSPC: European Randomized Study of Screening for Prostate Cancer
PLCO: Prostate, Lung, Colorectal, and Ovarian Cancer Screening Trial
PSA: Prostate Specific-Antigen
USPSTF: US Preventive Services Task Force

