## Supplemental Materials for "Potential impact of a population-based screening program on the increased burden of prostate cancer in Thailand: A simulation study"

**Supplementary Figure 1.** Weibull (dashed lines, model estimates) vs Kaplan-Meier (solid lines, empirical estimates) survival curves by stage for prostate cancer incidence in the Songkhla Cancer Registry.

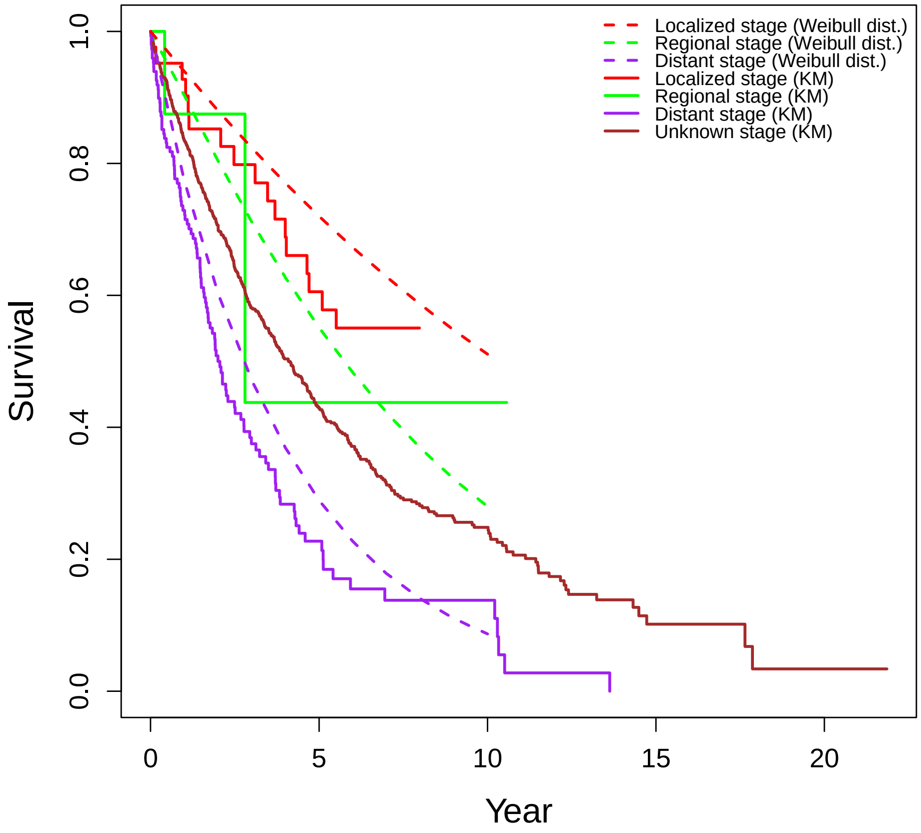

**Supplementary Figure 2.** Flowchart of the simulation analysis for PSA screening.

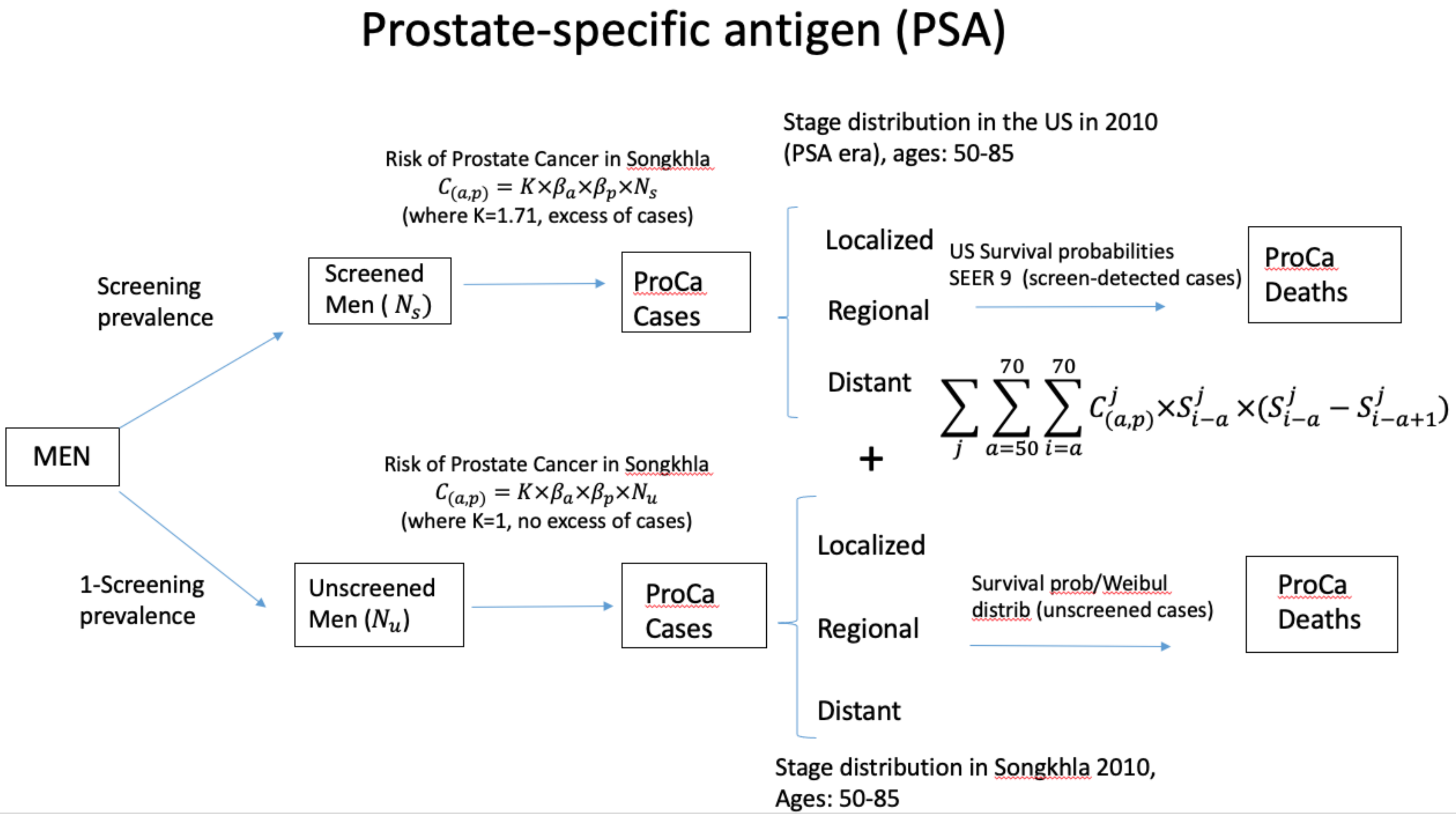


*K= 1.5 (lower bound) 2.0 (upper bound). ProCa Cases=Prostate cancer cases; ProCa Deaths=Prostate cancer deaths.

**Supplementary Table 1.** Cumulative number of prostate cancer cases and deaths (ages 50-70) under various screening uptake rates with three selected screening coverages (15%, 60%, 100%) in the Thai population.

| | | ERSPC | | | PLCO | | |
|---|---|---|---|---|---|---|---|
| | No Screen | 15% | 60% | 100% | 15% | 60% | 100% |
| Cases [1] | | | | | | | |
| Stage I & II | 194 | 1,168 | 4,085 | 6,685 | 1,282 | 4,534 | 7,433 |
| Stage III | 898 | 863 | 751 | 653 | 780 | 422 | 108 |
| Stage IV | 4,046 | 3,453 | 1,667 | 84 | 3,464 | 1,715 | 162 |
| Deaths[1] | | | | | | | |
| Stage I & II | 45 | 77 | 170 | 254 | 81 | 187 | 283 |
| Stage III | 289 | 252 | 140 | 42 | 247 | 119 | 7 |
| Stage IV | 1,821 | 1,555 | 754 | 44 | 1,561 | 779 | 86 |
| Cases[2] | | | | | | | |
| Stage I & II | 194 | 1,502 | 5,425 | 8,915 | 1,654 | 6,022 | 9,911 |
| Stage III | 898 | 896 | 880 | 874 | 784 | 443 | 143 |
| Stage IV | 4,046 | 3,456 | 1,686 | 115 | 3,473 | 1,747 | 214 |
| Deaths[2] | | | | | | | |
| Stage I & II | 45 | 90 | 221 | 339 | 95 | 243 | 377 |
| Stage III | 289 | 255 | 149 | 56 | 247 | 120 | 9 |
| Stage IV | 1,821 | 1,556 | 764 | 61 | 1,565 | 796 | 113 |

[1] 1.5 excess of cases. [2] 2.0 excess of cases
ERSPC=European Randomized Study of Screening for Prostate Cancer; PLCO=Prostate, Lung, Colorectal, and Ovarian (PLCO) Cancer Screening Trial.

**Supplementary Figure 3.** Prostate cancer stage distribution by different screening uptake rates (no screening vs. screening with uptake 15%, 60%, 100%) under the ERSPC (top panels) and PLCO (bottom panels) trial scenarios. Two different excess risk levels are used: 1.5 vs. 2.0. ERSPC=European Randomized Study of Screening for Prostate Cancer; PLCO=Prostate, Lung, Colorectal, and Ovarian (PLCO) Cancer Screening Trial.

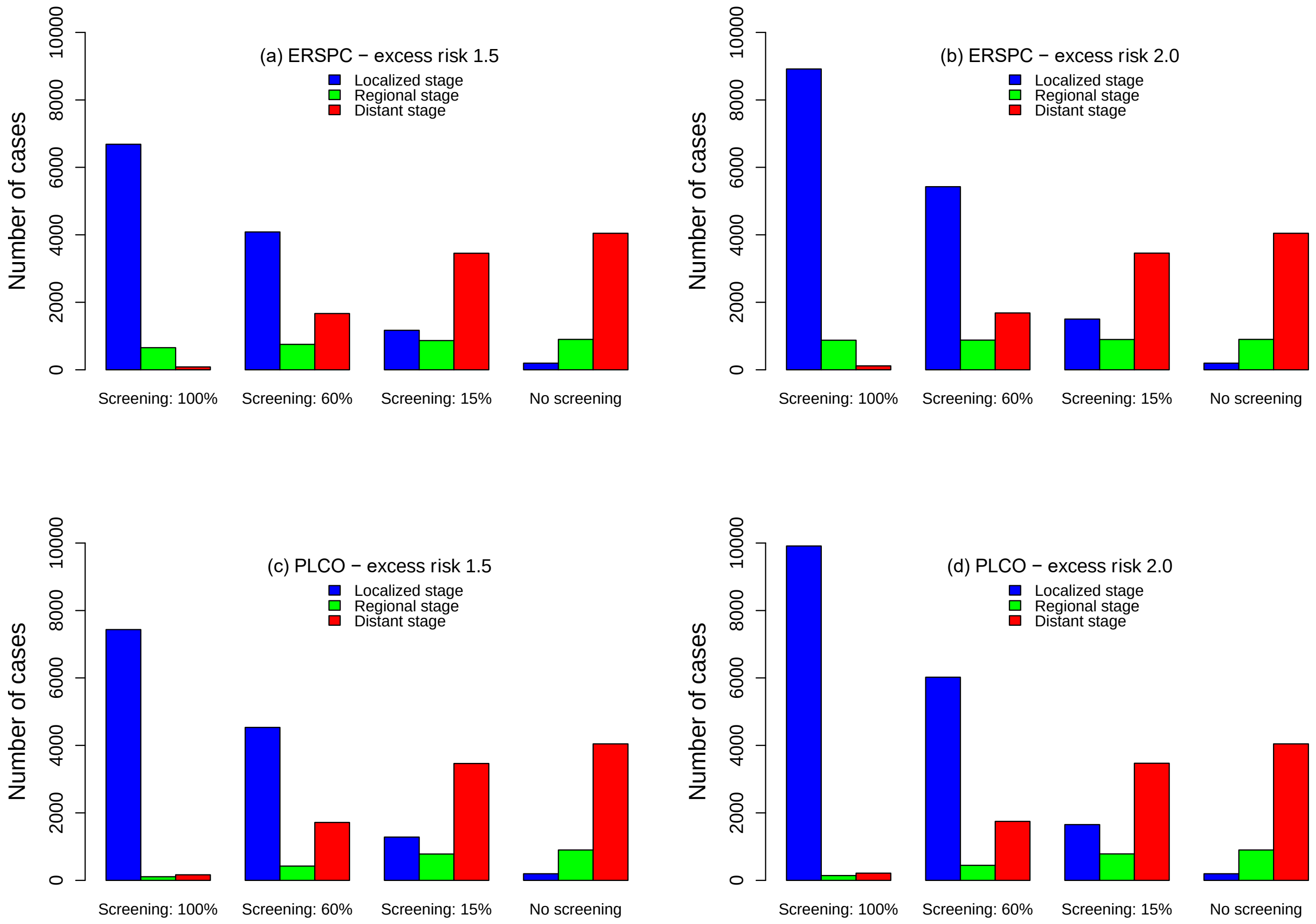

**Supplementary Figure 4.** Number of prostate cancer cases and deaths by different screening uptake rates (no screening vs. screening with uptake 15%, 60%, 100%) under the ERSPC trial scenario. Two different excess risk levels are used: 1.5 vs. 2.0. ERSPC=European Randomized Study of Screening for Prostate Cancer; PLCO=Prostate, Lung, Colorectal, and Ovarian (PLCO) Cancer Screening Trial.

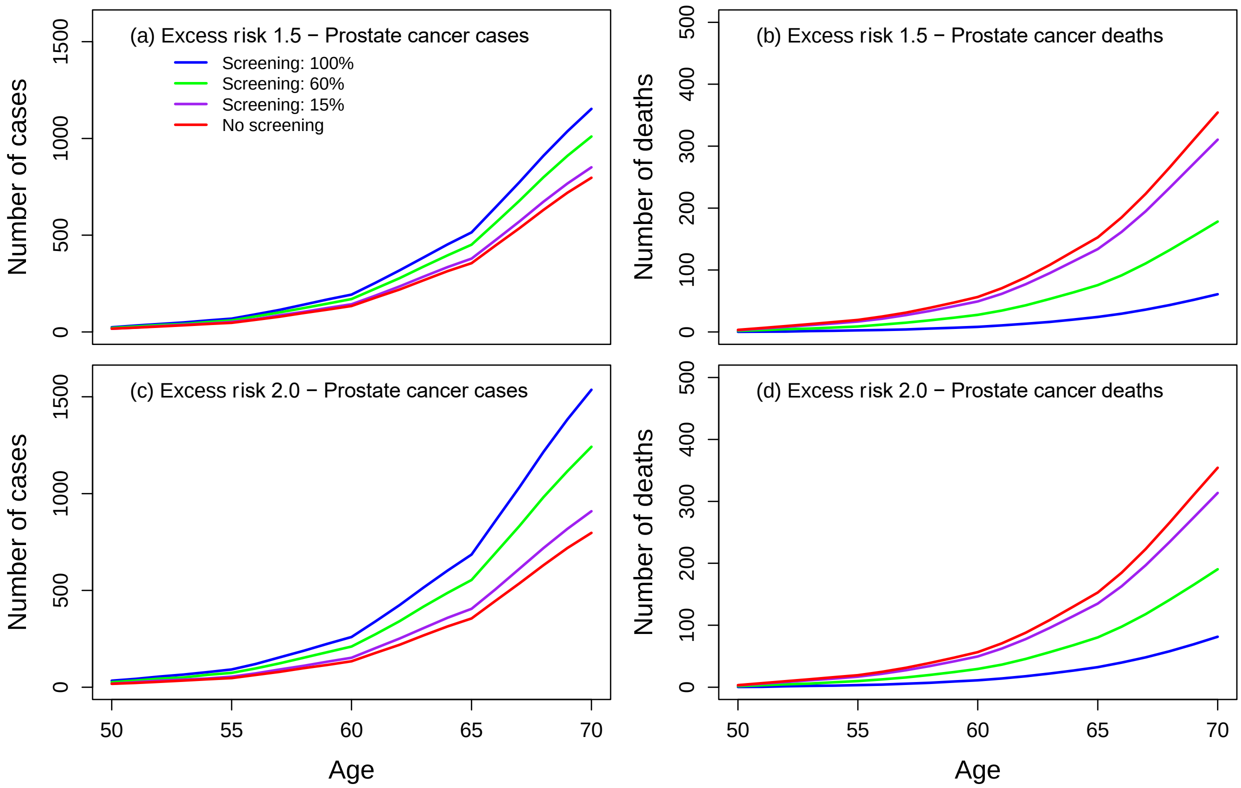

**Supplementary Figure 5.** Number of prostate cancer cases and deaths by different screening uptake rates (no screening vs. screening with uptake 15%, 60%, 100%) under the PLCO trial scenario. Two different excess risk levels are used: 1.5 vs. 2.0. ERSPC=European Randomized Study of Screening for Prostate Cancer; PLCO=Prostate, Lung, Colorectal, and Ovarian (PLCO) Cancer Screening Trial.

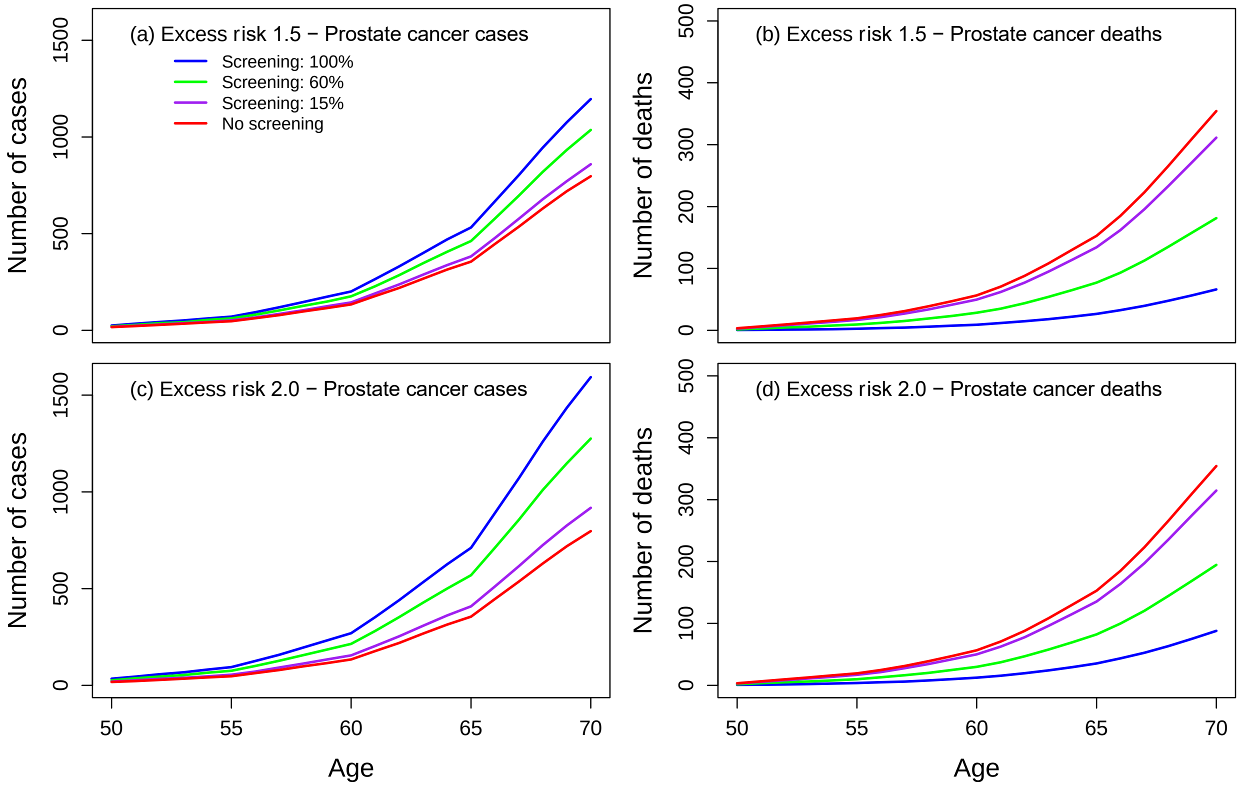

**Supplementary Figure 6.** Number of stage-specific prostate cancer cases by different screening uptake rates (no screening vs screening with uptake 15%, 60%, 100%) under the ERSPC trial scenario. Two different excess risk levels are used: 1.5 vs. 2.0. ERSPC=European Randomized Study of Screening for Prostate Cancer; PLCO=Prostate, Lung, Colorectal, and Ovarian (PLCO) Cancer Screening Trial.

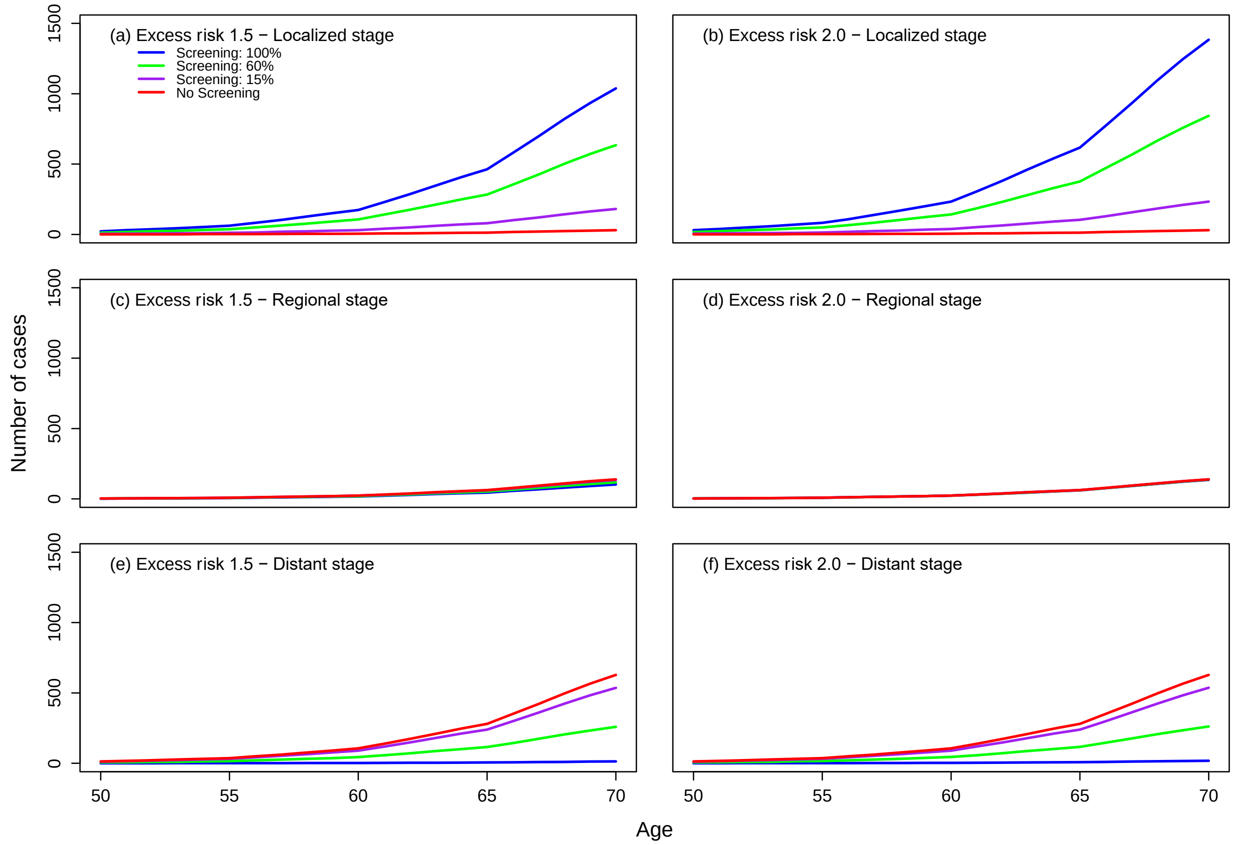

**Supplementary Figure 7.** Number of stage-specific prostate cancer cases by different screening uptake rates (no screening vs screening with uptake 15%, 60%, 100%) under the PLCO trial scenario. Two different excess risk levels are used: 1.5 vs. 2.0. ERSPC=European Randomized Study of Screening for Prostate Cancer; PLCO=Prostate, Lung, Colorectal, and Ovarian (PLCO) Cancer Screening Trial.

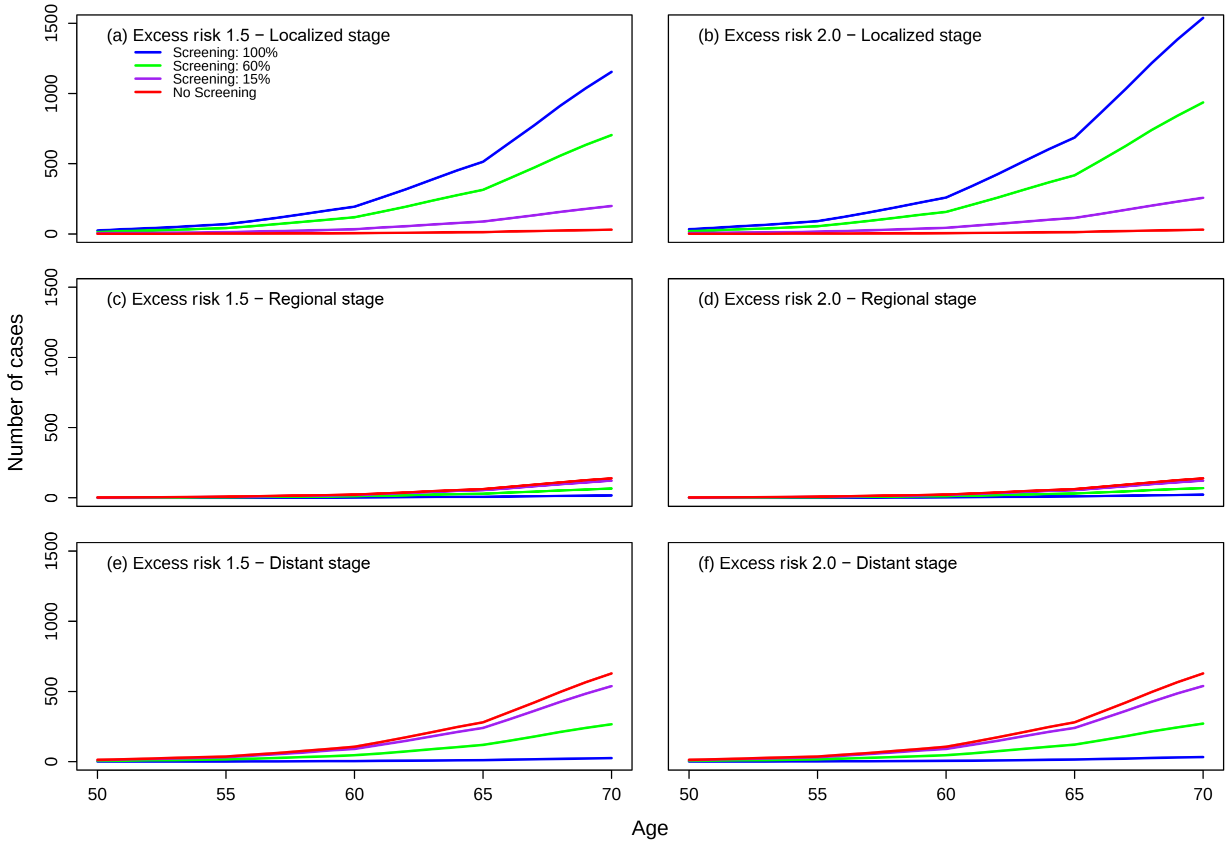

**Supplementary Table 2.** Case fatality ratio (CFR) under ERSPC and PLCO trial scenarios.

| | No overdiagnosis | | 23% overdiagnosis | | 42% overdiagnosis | |
|---|---|---|---|---|---|---|
| Screening uptake | ERSPC | PLCO | ERSPC | PLCO | ERSPC | PLCO |
| No screening | 0.42 | 0.42 | 0.42 | 0.42 | 0.42 | 0.42 |
| 15%[1] | 0.34 | 0.34 | 0.35 | 0.36 | 0.37 | 0.37 |
| 60%[1] | 0.16 | 0.16 | 0.19 | 0.18 | 0.21 | 0.21 |
| 100%[1] | 0.05 | 0.05 | 0.05 | 0.05 | 0.05 | 0.05 |
| 15%[2] | 0.32 | 0.32 | 0.34 | 0.34 | 0.35 | 0.36 |
| 60%[2] | 0.14 | 0.14 | 0.16 | 0.16 | 0.19 | 0.18 |
| 100%[2] | 0.05 | 0.05 | 0.05 | 0.05 | 0.05 | 0.05 |

[1] 1.5 excess of cases. [2] 2.0 excess of cases

ERSPC=European Randomized Study of Screening for Prostate Cancer; PLCO=Prostate, Lung, Colorectal, and Ovarian (PLCO) Cancer Screening Trial.
